# Exploration of barriers to providing mental health care and factors supporting access to mental health care in Australian prisons

**DOI:** 10.64898/2026.05.11.26352849

**Authors:** C. Comben, M. Burgess, Z. Rutherford, C. Meurk, L. Rivas, J. John, S. Diminic

## Abstract

**Objective:** This study aimed to understand barriers to providing mental health care in Australian prisons and explore factors supporting access to mental health care.

**Methods:** This qualitative study used focus groups with people with a lived experience of mental illness in prison or caring for someone in prison with mental illness and people involved in providing mental health care in prisons. Focus group transcripts were thematically analysed.

**Results:** Focus groups were held with eight participants. Identified barriers to providing and accessing mental health care in prison included perceived stigma, insufficient resourcing, logistics driven care, inconsistent standards, and tension between prison- and health-centred systems. Factors supporting access to mental health care in prisons included equivalence of care, individually tailored care, connection, resourcing, and coordinated care.

**Conclusions:** Identified barriers make providing mental health care in prisons difficult, resulting in unmet needs. Factors that support access highlight mechanisms available to improve outcomes, but their utility depends on addressing all barriers.

**Implications for Public Health:** Identified barriers and supporting factors can help guide service design to improve access and promote interagency collaboration across justice and health. Identified barriers can also be used to help inform evidence-based policy making, including workforce development and increased funding.

## Introduction

Access to adequate mental health care in Australian prisons is limited (1). People in prison are less likely to have their mental health needs recognised and responded to with an appropriate level of care and support (2). Despite more than half of people in prison having a current mental illness (3), only a small proportion access mental health care in prison (4) or are referred to community mental services on release (4). Out of necessity, limited resources in prison are typically oriented toward people with severe mental illness; people with less intensive or urgent needs do not receive adequate care (5), further exacerbating inequities. This limited access to mental health care in Australian prisons is likely explained by several factors, including exclusion from Medicare (5) and limited resourcing of prison mental health services, particularly in terms of available workforce (6).

Additional barriers to accessing mental health care in prisons have been identified internationally. Individual level barriers to accessing mental health care include perceived low quality of care, long wait times, difficulty in navigating the system, distrust of system, and stigma from both correctional staff and other incarcerated people (7-9). Interpersonal level barriers include corrections staff not responding appropriately to signs of distress and requests for help (7). Organisational level barriers include inadequate mental health training for all staff in prisons and low quality services (10). Finally, policy level barriers include lack of resources, including workforce (10). Despite these barriers, factors supporting access to mental health care in prisons have been identified, including strong relationships with peers and prison staff (7, 8).

Australian research to date has focused on barriers to accessing and delivering services within broader prison healthcare. Previous studies identified barriers caused by the misalignment of priorities and power differentials between corrections and health systems (11) and challenges with service navigation and loss of autonomy (12). Developing an understanding of barriers to accessing and providing mental health care in Australian prisons, and factors that support access, is important given the significant unmet mental healthcare needs in prisons and known fragmentation between the country’s health and justice sectors. This information provides important context for health service planning by helping to identify and address gaps between service benchmarks and current service provision. The aim of this study was therefore to understand the barriers to providing mental health care in Australian prisons and explore factors supporting access to mental health care.

## Methods

Focus groups were conducted to identify the mental health service needs of people in prison as part of a larger study developing inputs for a needs-based forensic mental health service planning model in Australia. Ethics approval was obtained from the UQ Human Research Ethics Committee (2021/HE002377). Reporting adheres to the Consolidated Criteria for Reporting Qualitative Research (COREQ) guidelines (13). CC, MB, ZR, CM, and SD are academic researchers with backgrounds in public health, epidemiology, mental health services research and evaluation.

### Participants and recruitment

Participation was sought from people with lived experience of mental illness in prison, carers, and stakeholders from prison and forensic mental health and health services, corrective services, and non-government organisations involved in transitional care.

An expression of interest form was sent to an Australian network of senior forensic mental health clinicians and relevant organisations, including professional and peak bodies, with requests to disseminate through their networks.

A selection matrix considering role, jurisdiction and availability was used to select prospective participants for focus groups, prioritising lived experience participants. To support safety and anonymity, individuals from mental health services from the same jurisdiction as any lived experience participants were not included within the same focus group.

### Data collection

Three online, repeated focus groups were facilitated by CC, SD and JJ in February 2022. Participants were asked about (1) the factors that drive individual needs for mental health services in prisons and (2) the mental health service mix required for people with different severities of mental illness in prison, using a semi-structured guide (see Appendix I). Sessions were recorded and transcribed verbatim by an external transcription service and de-identified by the research team.

### Analysis

Data were analysed using a descriptive thematic analysis (14, 15). Two authors (CC and MB) read and coded all anonymised transcripts using an inductive approach to identify barriers to providing mental health care in Australian prisons and factors supporting access to mental health care using NVivo 14 (16). Initial coding was refined through an iterative, collaborative process. Coding disagreements were discussed and resolved through discussion. Coded data were organised by shared meanings to generate themes and sub-themes. A thematic map was created to align themes with the spheres of influence within a socioecological model (17, 18). This model was used to identify leverage points that could affect change on the ability to provide good mental health care in prisons. At the individual level, themes were developed on the assumption that people in prisons are willing and able to engage with services.

To ensure analytic rigour, all co-authors contributed to different stages of the study. The lead author met regularly with the senior author. Other authors provided critical input to the analysis and manuscript.

## Results

Ten participants were invited, and eight participants from four jurisdictions (Queensland, New South Wales, Victoria, Australian Capital Territory) took part (see Appendix II). Three had lived experience of mental illness in prison or caring for someone in prison with mental illness. Five worked in prison mental health services as service directors, managers, or direct care providers.

Ten themes and twelve subthemes were identified. Five themes related to barriers to providing and accessing mental health care: perceived stigma, insufficient resourcing, logistics driven care, inconsistent standards, and tension between prison- and health-centred systems (Figure 1). Five themes related to factors supporting access to mental health care in prisons: equivalence of care, individually tailored care, connection, resourcing, and coordinated care (Figure 2). Barriers were identified at the individual, organisational and policy levels of the socioecological model, while factors supporting access to mental health care in prisons were identified at the interpersonal, organisational and policy levels (Figure 3).

**Figure 1.**
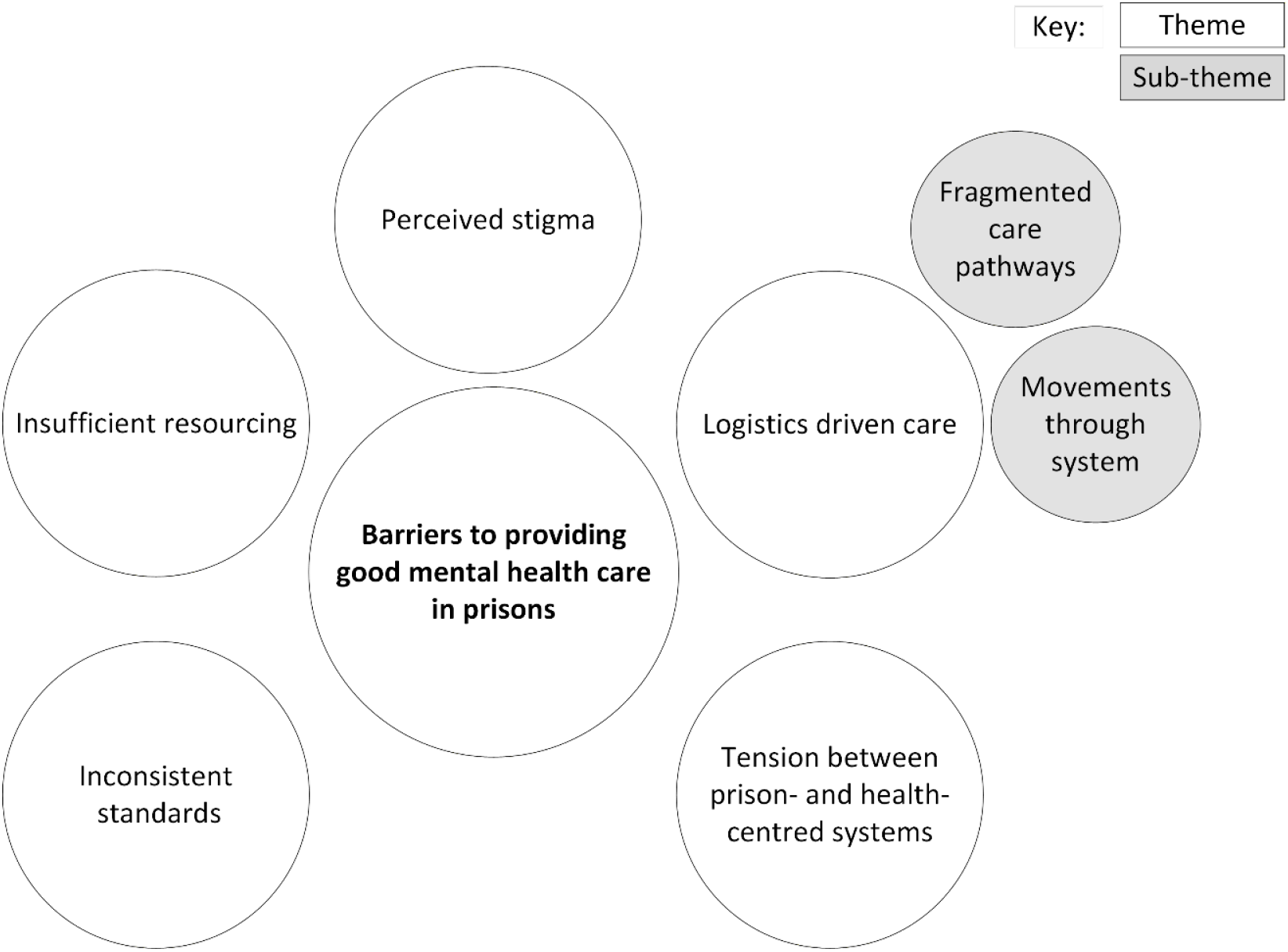
Thematic map of barriers

**Figure 2.**
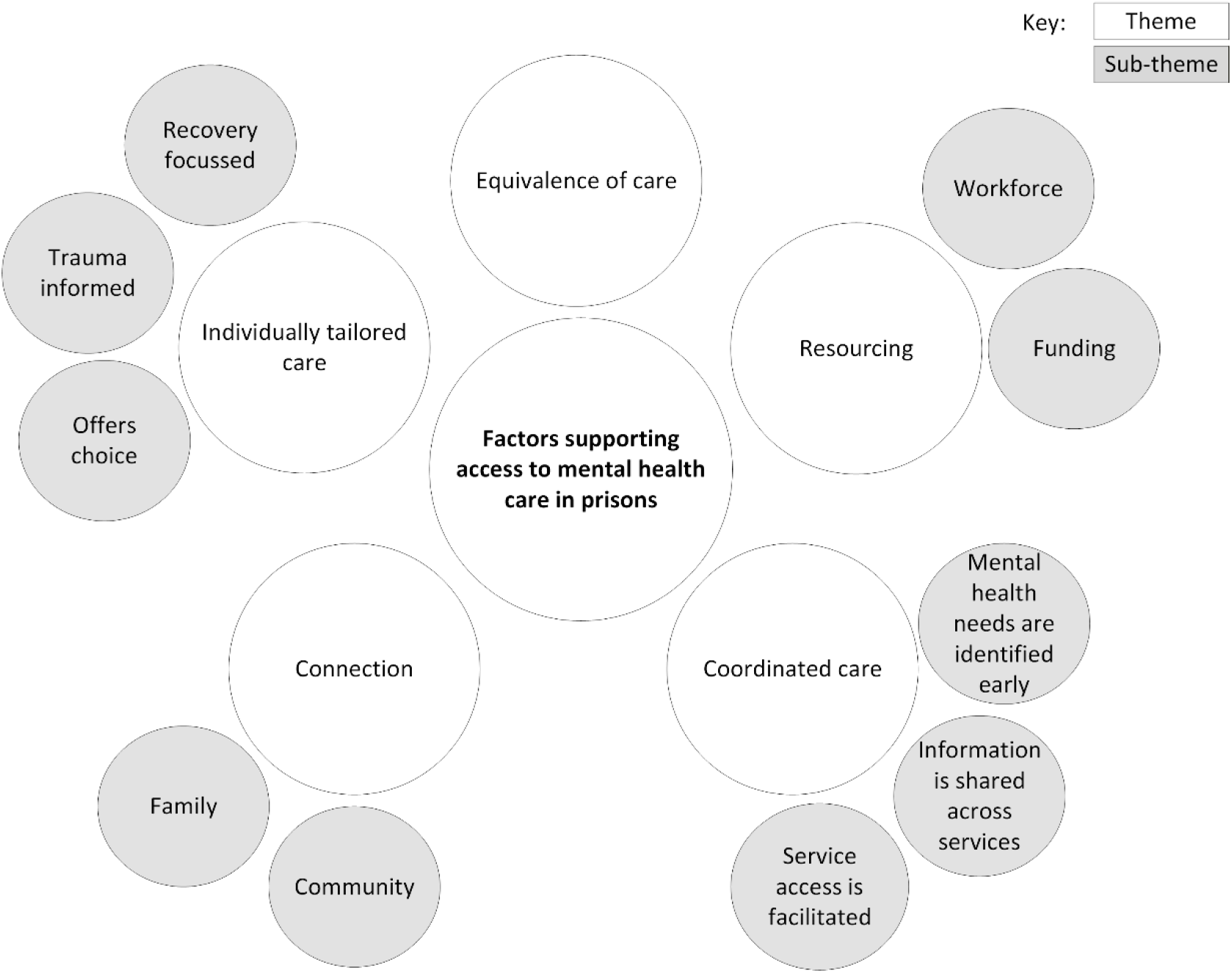
Thematic map of factors supporting access to mental health care in prisons

**Figure 3.**
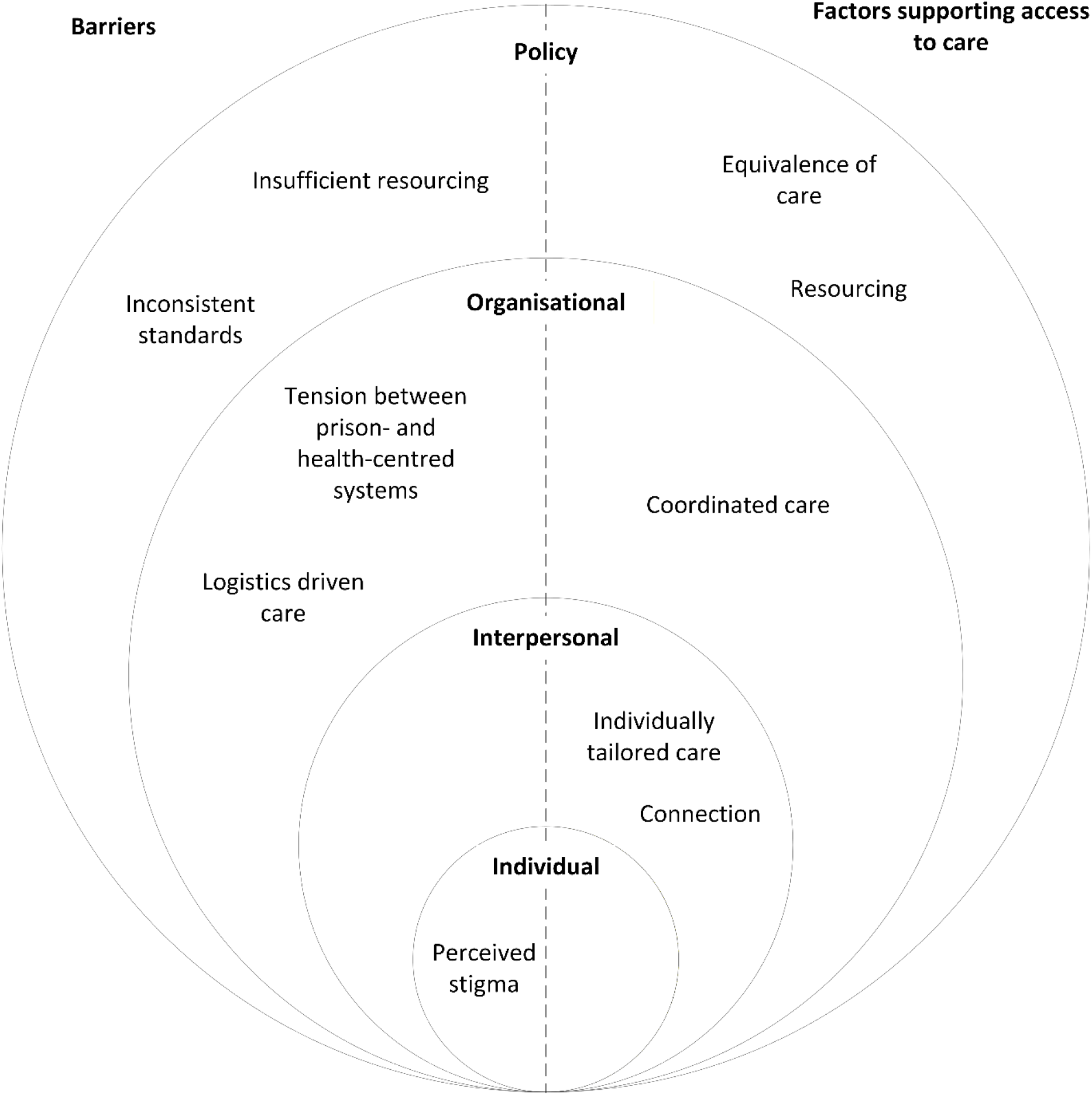
Overlay of socioecological model with identified barriers and factors supporting access to mental health care in prisons

### Individual level

#### Barriers

At the individual level, perceived stigma was described as a barrier to accessing mental health care in prison.

> *“There’s that sort of dirtiness that comes with being in a criminal justice system and being labelled as an offender”* (Participant 6)

This extended to preferring not to access mental health care while in prison due to stigma.

> *“There is that stigma where someone doesn’t want to be in the psych unit, they don’t want to be in the protection unit, they want to just do their time”* (Participant 3)

Another participant noted concerns about repercussions of accessing care:

> *“Usually the main problem for anybody in prison, […] the person who’s in, is afraid of repercussions if they make a stink about anything.”* (Participant 1)

### Interpersonal level

#### Factors supporting access to care

##### Individually tailored care

Participants described several principles to guide care delivery and therapeutic relationships, including care that is recovery focussed, trauma informed and offers choice. Choice in receiving care was reiterated, as people in prison may prefer to stay under the radar to avoid being seen as a problem by peers and staff. As such, the need to *“empower individuals to seek assistance as they wish and as they need it”* (Participant 5) was described.

Recovery-focussed care was also identified as a key principle, especially in the context of providing perspective and hope for the future.

> *“We can’t undervalue how important […] giving people hope, giving them perspective, it’s a recovery – […] focussed way of doing things”* (Participant 1)

One participant with lived experience described the importance of recovery-focussed care:

> *“That would be invaluable to show look, there is life beyond this, there are options beyond this”* (Participant 3)

Relating to trauma informed care, participants raised the need to recognise and respond to the inherent trauma of prisons. One participant said:

> *“Being put in a prison, especially if you’ve never been there before or you have a degree of mental illness, it has to be one of the stressful things that could happen to you in this country.”* (Participant 3)

##### Another participant reiterated

> *“The very place of prison and adjusting to that change to somebody’s life, which is pretty drastic is trauma, it’s traumatic. Even if that person doesn’t experience direct physical trauma or coercion from anybody else, just witnessing on the day-to-day basis, the behaviour of others, be it towards the guards or from the guards or towards each other, is extremely traumatic.”* (Participant 1)

Finally, participants identified a need for cultural responsivity in services, describing *“there is not any real cultural responsivity built into our systems of prison. In our heads I think we’ve designed them for many white young males and that’s about it.”* (Participant 6)

##### Connection

Participants emphasised supporting people in prison to maintain connections with their families and communities. The caring role was highlighted, with support for carers needed while in custody and in preparation for release. One participant described the types of family supports available in their jurisdiction as “*the liaison connecting person between the person who’s in the prison and the families on the outside”* (Participant 1).

The same participant elaborated on the types of supports needed to help prepare carers to reunite with their family member after their release from custody:

> *“how to prepare and cope when the person’s going to be on leave or come home, how to help the family members cope.”* (Participant 1)

Additionally, many people in custody may have been carers themselves prior to their incarceration, such as for young children. Separation of people in custody from their own supporters or carers was also raised, especially in the context that carer voices are marginalised for adult populations due to the emphasis on person-centred care:

> *“The emphasis on being person centred assumes that the person is an individual entity operating without anything else in their life”* (Participant 1)

As such, participants advocated for open communication pathways between people in custody and their families and carers. Even while their family member is in prison, participants suggested that family are often best placed to give insight into the person’s current mental health state:

> *“Often it’s the family who can give the best insight even if they’re on the outside because they’re the ones having the weekly phone contact.”* (Participant 1)

Participants also stated that when people in prison are displaying stress signs, additional support or contact from family may be helpful to try and prevent further escalation. In addition to families, participants explained the importance of maintaining connections with the broader community while in custody. Connections in the community often serve as the links for the person when they are released and ideally support them to prevent reincarceration. Most people in prison do not have good supports in the community, highlighting the need to bolster these supports while they are in custody to enhance outcomes post-release.

### Organisational level

#### Barriers

##### Logistics driven care

Participants described challenges associated with the logistics-driven nature of prisons. Fragmented care pathways were highlighted as one barrier. Specific challenges affecting continuity of mental health care included siloed approaches, differing systems and differing principles. The siloed operations of the various systems within prisons were a key issue described by participants. One participant said:

> *“Being recepted into custody is awful for so many reasons but one of them is that you […] end up on this […] conveyor belt of retelling your story and reassessment at multiple different points”* (Participant 4)

##### Another participant said

> *“lots of agencies are operating as individuals and we don’t necessarily talk to each other literally or figuratively.”* (Participant 5)

##### Another participant said

> *“We currently have […] multi agency arrangements that are very fragmented […] and that service and systems fragmentation means there’s lots of points of gaps where people can fall through and do very often”* (Participant 6)

Participants described challenges in accessing and providing prison mental health care due to the logistical movements of people in custody. These movements create operational challenges, such as visiting healthcare providers or lawyers unable to locate their clients to provide services, and clinical challenges, as an individual’s physical location may define the services that are available to them. Participants also described the traumatic nature of logistical movements. One participant said:

> *“People being moved from facility to facility is exceptionally traumatic for them. They’re generally not given any notice or much notice, they have absolutely no say in it, it drastically affects the family and visitation or connection with what’s meaningful to them. Sometimes it can have drastic impacts on them physically.”* (Participant 1)

##### Tension between health- and prison-centred models

At the organisational level, tension between corrective and health services within prisons was emphasised as a key barrier to providing good mental health care. One reason for this tension is the fundamental difference in the philosophies of each organisation. One participant stated:

> *“It’s not really seen as a care industry, it’s seen as a security industry. So you’ve got two really competing philosophies banging into each other in the forensic mental health system.”* (Participant 6)

Participants also described differences in the factors that input into decision making within each organisation. Factors important to corrective services tend to be related to logistics, such as “where is there enough room for this person tonight?” and “where is the magistrate sitting tomorrow?”. Conversely, the health-centred organisations consider factors such as:

> *“How unwell is this person, how much care does this person need, where are their family. Those humanist elements are not primary in the [corrective] service system.”* (Participant 6)

Participants stated these strengths-based considerations are *“not the language or the thinking that’s used in a punitive system”* (Participant 1). One participant stated that within correctional services, there’s *“no shame in using […] very stigmatising terms”*, highlighting:

> *“In fact, it’s called an offender management pathway or a framework. So all the literature that correctional officers are working from and officers is another good example of the type of language.”* (Participant 6)

One participant said that mental health services are not uncommonly stigmatised by corrective services:

> *“The organisation I work for actually had some of our staff go into [prison] to provide classes and to try and prepare people for the outside. […] they weren’t welcomed all that much.”* (Participant 3)

Additionally, there is often a lack of cooperation between the two organisations, contributing to the complexity of providing mental health care in prisons. One participant said:

> *“[The] health team makes recommendations to corrective services about location and accommodation, even within the prison. […] recommendations are made but not necessarily followed.”* (Participant 2)

##### Another participant explained

> *“We can put a flag to say hey, if you’re going to transfer this person can you let us know in advance, so that we can do some prep around it. But […] they make the decision and we react.”* (Participant 4)

#### Factors supporting access to care

##### Coordinated care

Participants highlighted the importance of coordinated care at the organisational level. First, they described the importance of facilitating service access, especially when people first come into prison.

##### One participant said

> *“A lot of it is providing that education and orientation when they come into prison […] this system is really very difficult to navigate but we’ve tried to orientate people when they come in.”* (Participant 5)

Participants also described the importance of linking people to services that continue in the community after release.

Secondly, early identification of mental health needs was emphasised. Participants suggested reception screening should be conducted by specialist prison mental health services to reduce the number of people whose needs go undetected.

> *“I think then we’d have hopefully less people that fall through the gaps that rely on referral pathways from other services.”* (Participant 4)

In the context of assessment, participants reiterated the importance of considering collateral information from families and carers where available, stating:

> *“[Family] know the person well enough, they can tell by their tone of voice where things are at by what’s not said as well as what is said or from letters that they might receive or anything they’re – when they’re able to visit from visits. They will get a perspective that might even be truer than going to someone’s office or a clinical visit.”* (Participant 1)

Participants reiterated the importance of information sharing pathways within and across services. Transfers between prisons for logistical reasons, often with little notice, interrupt continuity of care. A regional coordinator may help support this continuity of care:

> *“People may have had appointments scheduled or care needs in one location when they got transferred and then that all fell by the wayside. […] Having a coordinator who can oversee and ensure that people aren’t lost, they don’t fall through the cracks and that communication between different locations occurs, I think that’s probably a lot better use of resources and more helpful, really, across the whole system.”* (Participant 5)

Participants also described the importance of comprehensive transfer of care. In the context of transfers across prisons, one participant stated:

> *“You’d want to see that there’s a prison mental health team that sits in each prison and […] they work in a coordinated way to be able to pick that up in a process that enables that good communication. So when someone moves from one prison to another […] the next prison mental health team just picks them up, just like a transfer of care within the community.”* (Participant 4)

### Policy level

#### Barriers

##### Insufficient resourcing

At the policy level, insufficient resourcing for prison mental health care was a common theme. Participants described challenges in providing best practice care due to limited resources, explaining that they are *“competing for a very small pot of money […] with a lot of other services”* (Participant 5). One specific barrier was the lack of beds for transfers to secure treatment orders:

> *“I always wonder how many we don’t actually recommend for [secure treatment orders] because we kind of know that they’re not going to get into the hospital anyway. […] Some of these people are waiting in excess of 80 days, so they never actually get there. They get released before they get into hospital which is pretty poor of us really when you think if they were having a heart attack or were in ED they wouldn’t just be left to wait for the bed. So, it’s pretty dire.”* (Participant 5)

Other key issues included lack of resourcing for cultural supports such as Indigenous mental health workers and the gap in psychological intervention for people with mild levels of mental illness created by the Medicare exclusion. One participant elaborated further on the Medicare exclusion:

> *“What does exist as a gap is the psychological intervention or therapeutic intervention that you would get in that community setting where you have a mild mental health concern where you have access to Medicare and therefore mental health plans and that not, Medicare, not existing in the custodial space is a huge impediment.”* (Participant 4)

##### Inconsistent standards

A second barrier at the policy level was the inconsistent standards across Australian jurisdictions in terms of prison health, mental health and corrective services models. One participant explained:

> *“State and territory differences really can’t be ignored because they shape everything in terms of your experience of going through these systems and working in these systems. I’ve worked in a couple of different states so I think it does take time to understand and to navigate it.”* (Participant 6)

These inconsistences were in part attributed to different legislation, especially regarding involuntary mental health treatment in prisons. Another key inconsistency highlighted was the different ways of assessing need for mental health care across jurisdictions, including use of different assessment tools.

##### One participant stated

> *“Perhaps we are lacking […] a coordinated national way of assessing the need. […] I think we all develop our own methods and assessment tools by the sounds of it.”* (Participant 5)

#### Factors supporting access to care

##### Equivalence of care

At the policy level, the principle of equivalence of care was a factor supporting access to mental health care in prisons. Participants described how service models have been designed to emulate community equivalents and offer comprehensiveness of services. One participant explained the application of this factor in practice:

> *“Mild [mental illness treatment] would usually [be provided] in the custodial space – their mental healthcare would be managed by our primary healthcare services around that equivalence of care to community, with GP type care.”* (Participant 4)

However, there was variety in the models described. One participant explained that in their jurisdiction, it was still a mental health-specific response provided at the primary care level:

> *“I think it’s appropriate that we do have primary mental health. It’s not just primary care, but the primary mental health clinicians, they have [Registered Psychiatric Nurses] there. It’s just that if there’s a tertiary need for a service then [specialist mental health] gets involved. I think it’s a pretty good system because it is consistent with the community in virtually the same way as you’d see a GP first before going up to a specialist.”* (Participant 5)

One participant suggested that rather than focussing on equivalence of care, the focus should be on equivalence of outcomes:

> *“The prison population isn’t meant to have additional services beyond what the community would receive. But given the circumstances, that just seems entirely inadequate because part of the reason they’re in prison often is because of their mental health situation.”* (Participant 2)

##### Resourcing

Participants stated that advocating to local ministers and justice organisations for additional resourcing for forensic mental health was a key part of their role. One participant explained:

> *“We have to kind of put business cases together and really advocate why we need certain services. So I think we do need to be a bit strategic if we’re thinking nationally what we should be funding.”* (Participant 5)

Participants also described the need to consider workforce capacity, capability and training to support qualified staffing. One participant stated:

> *“What should we be aiming at so that there is adequate training, adequate tertiary positions, adequate workforce.”* (Participant 1)

The need to consider minimum qualifications and standards for people working in corrections was also raised:

> *“If you’re going to be based in correctional systems and settings you’re only as good as that service system as well. So that service system needs to have a light shone on it too. Correctional officers across Australia nationally, there is no minimum standard or qualifications.”* (Participant 6)

## Discussion

This study has identified barriers to providing mental health care in Australian prisons and factors supporting access to care in this setting. Barriers and factors supporting access to care were identified across all levels of a socioecological model, highlighting the complex and interrelated influences affecting provision of this care. This study found that many of the barriers to providing mental health care in prisons internationally are applicable to the Australian context, including inadequate funding, perceived stigma, limited workforce, fragmented care pathways and the prison setting itself (7-10, 19). Additional barriers unique to the Australian context were identified, including the inconsistent standards for services created by jurisdictional differences in legislation and screening tools, the siloed approach to service provision that causes fragmented care pathways including lack of transition planning, and the contribution of the Medicare exclusion to lack of service access.

Broadly speaking, the identified factors supporting access to mental health care in prisons align with established principles in general and forensic mental health care (20-22), such as equivalence of care, individually tailored care and coordinated care. However, these factors are undermined by the identified barriers. For example, delivering care that is equivalent to what is offered to community populations requires sufficient service resourcing and workforce.

A key finding is that there is insufficient resourcing currently allocated to the provision of mental health care to people in prison. While there are workforce shortages across most health service settings in Australia, they are especially pronounced in prisons, limiting access to care (1, 6). These workforce shortages may be exacerbated by difficulties in attracting healthcare professionals to work in corrections (23). For health professionals, there are barriers preventing employment in this space (24), especially for First Nations workforce (25). Advocacy for sufficient resourcing was raised as a factor supporting access to care by participants, including workforce planning to ensure qualified workforce are available when needed. More pathways are needed build a capable workforce, including entry level recruitment with support to grow and engage in professional development activities.

A second key barrier is the inconsistency in standards across Australian jurisdictions, with each operating under its own structure, resourcing and logistical arrangements. This is a known challenge, as there are no national standards in the provision of mental health care to prison populations (26). In particular, the lack of standardised assessment tools limits the ability to assess equity of care and track progress nationally. Conducting mental health needs assessments in a nationally consistent way may help to improve equity in mental health care by ensuring the same quality of assessment practices are available nationally. It may also lead to better outcomes for people requiring mental health care in prisons by enabling systematic data collection to inform resource allocation and policy.

Tension between health and prison-centred models of care was also identified as a barrier. Although previously identified as a barrier in other areas of prison health care in Australia (11), this study was able to explore this barrier as it relates to mental health care. The nature of prisons, which prioritise order, routine and control is at odds with safe, therapeutic environments for providing mental health care (1). For example, correctional health services can stigmatise mental health concerns, and recommendations about a person’s living requirements within a prison are not necessarily followed. These challenges highlight the need to consider correctional officers as a rehabilitation-oriented workforce to promote the rehabilitative role of officers above the traditional custodial, punishment and surveillance roles (27). Shifting the ethos of correctional services from a custodial focus to a rehabilitation focus will help to address the barriers created by logistics-driven care and could also provide additional benefits such as the increased well-being of individuals within prisons and better outcomes after release (28). Additionally, the tension between prison- and health-centred systems makes information sharing across different teams and services challenging, requiring individuals to advocate for themselves to ensure they can access adequate care. However, this is often problematic due to the perceived stigma associated with needing mental health care. Improving the cooperation and communication between health and correctional systems may therefore also help to improve access to mental health care in prisons.

While many of the factors that support access to mental health care in prisons offer clear and actionable recommendations for improving services, their implementation is challenging due to the various barriers identified in this study. For example, equivalence of care was also raised as a key factor supporting access to mental health care in prisons. While equivalence of care is stated as a priority in policy documents both in Australia and internationally (20, 29), this study highlights that is not happening in prison mental health care due to a range of systemic barriers and a lack of resourcing, including the exclusion from Medicare. The inability to provide this equivalence of care breaches human rights and ultimately impacts on the outcomes of people in prison, such as increased recidivism due to poorer mental health while in prison (29, 30).

## Limitations

There are several limitations of this study. First, the participants in this study were not as diverse as originally intended. Initial recruitment sought participants from prison health services (i.e., primary care), corrective services, and non-government organisations who interface with the transition from custody to the community. All service-related participants in this study were from prison mental health services. The perspectives of other stakeholder types involved in the provision of mental health care to people in prison may have been different. Furthermore, there were no participants from Tasmania, Northern Territory, South Australia or Western Australia. The perspectives of participants from these jurisdictions may have been different to those from Australia’s most populous and urban jurisdictions, who were represented in this study.

Second, few participants had a lived experience of engaging with mental health care in prisons, therefore the barriers and facilitators identified largely reflect service provider perspectives. This limits understanding of individual barriers to help-seeking in prisons. However, this study aimed to generate information to support health service planning, making the service provider focus beneficial for understanding internal system improvements.

Finally, participant biases and motivations may have influenced participation and responses. Participants were recruited into a study seeking to enable needs-based mental health service planning by generating profiles of what good mental health care looks like within Australia’s forensic mental health system. Therefore, participants were likely already engaged in system development activities and seeking avenues through which they could contribute to the improvement of prison mental health care. Due to their role and engagement in service improvement, their opinions may have been different to other mental health service stakeholders in prisons.

## Implications

The barriers identified in this study make providing good mental health care in prisons difficult, resulting in unmet needs. Factors that support access highlight mechanisms available to improve outcomes, but their utility depends on addressing all barriers. At a policy level, barriers can inform evidence-based policymaking, including workforce development and increased funding. Needs-based planning of mental health services in prison can enable quantification of workforce and resourcing shortages. At the organisational level, barriers and supporting factors can guide service design to improve access and promote interagency collaboration across justice and health. Ultimately, it is critical to support access to care that is integrated, well-resourced and continuous across the criminal justice system, improving outcomes for people in prison and after release.

## Data Availability

The data generated in this study are not publicly available due to ethical and confidentiality considerations.

## Statements and declarations

### Funding

This paper was derived from original research conducted for the National Mental Health Service Planning Framework project funded by the Australian Institute of Health and Welfare and Department of Health, Disability and Ageing. This publication reflects the views of the authors and should not be construed to represent the views or policies of the Australian Institute of Health and Welfare or the Department of Health, Disability and Ageing.

### Ethics

The Human Research Ethics Committee of The University of Queensland gave ethical approval for this work (2021/HE002377).

### Conflicts of interest

The authors declared no potential conflicts of interest with respect to the research, authorship, and/or publication of this article.

## Appendix I

### Indicative focus group questions

1. Drawing on your experiences with prison populations, and thinking of people who have a mild/moderate/severe mental illness in that setting, what are the factors that contribute to/drive an individual’s need for mental health services while they are in prison?
2. Based on the factors we have just discussed, and drawing on your experiences, do you think there are any distinct sub-groups of people in prison with mild/moderate/severe mental illness who have a similar intensity and mix of mental health service needs?
3. The following care profile from the NMHSPF is for people with mild/moderate/severe mental illness. Drawing on your experience, do you think this mix of services is correct for people in prisons? (display care profile on screen)
  a. Are any services missing?
  b. Are any services inappropriate?
  c. Are the proportions of the need group requiring the service appropriate?
  d. Are the providers appropriate?
  e. Is the amount of services included appropriate?

**Figure A1.**
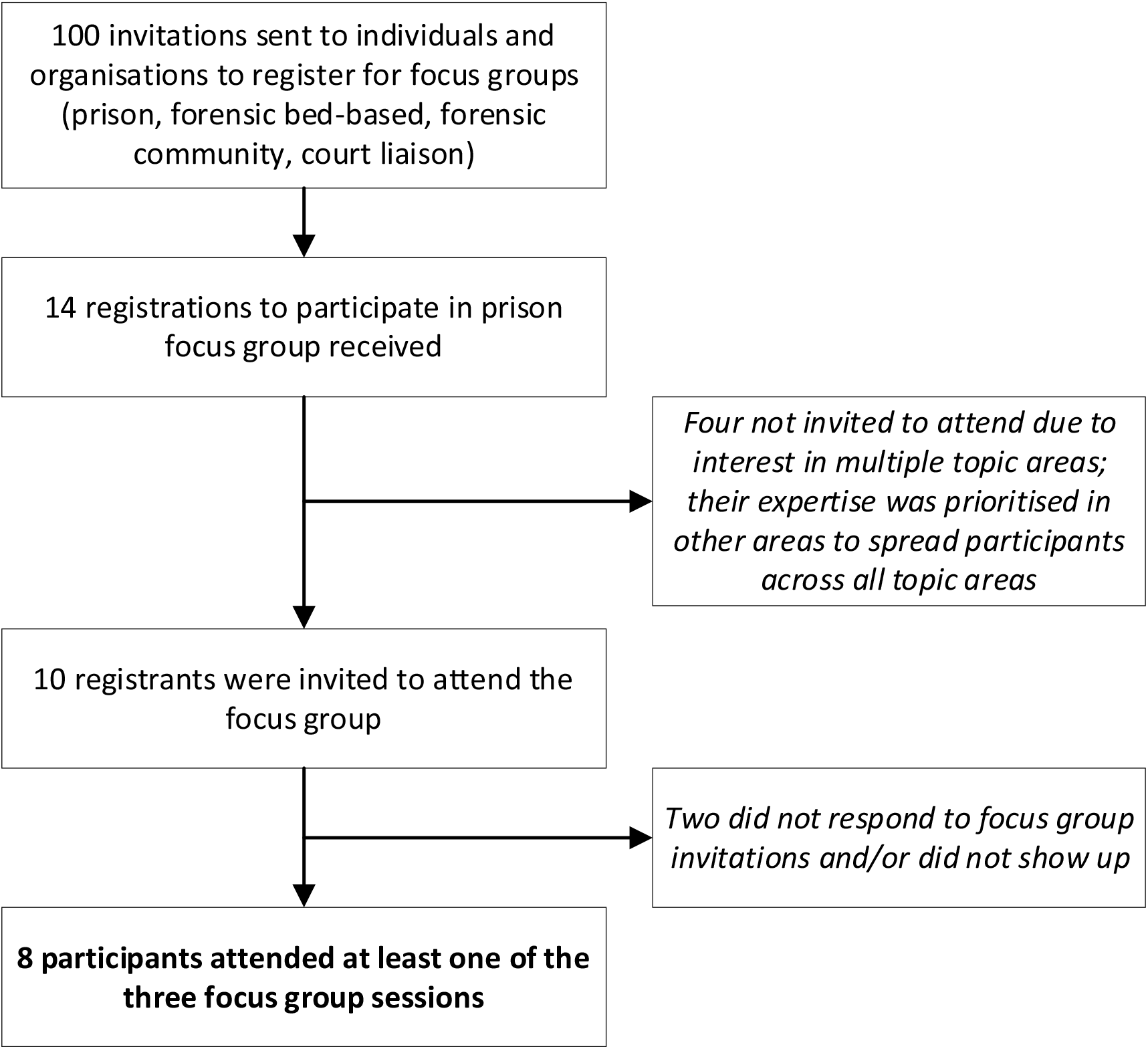
Focus group recruitment flowchart

## Notes

### Competing Interest Statement

The authors have declared no competing interest.

